# A multinational e-survey on the delivery of cardiology services in Africa during the COVID-19 pandemic: what should we expect after this pandemic?

**DOI:** 10.1101/2020.07.24.20160390

**Authors:** Chris Nadege Nganou-Gnindjio, Mazou Ngou Temgoua, Liliane Mfeukeu Kuate, Clovis Nkoke, Joel Noutakdie Tochie, Valerie Ndobo-Kue, Amalia Owona-Nsiaguam, Jerome Boombhi, Richie Kipenge, Urbain Huba, Malick Kane, Mohamed Taha el Jirari, Sylvie Ndongo-Amougou, Bâ Hamadou, Alain Menanga, Samuel Kingue

**Author notes:** Corresponding author: Dr Mazou Ngou Temgoua, MD, MPH, Faculty of Medicine and Biomedical Sciences, Department of Medicine and specialities, Yaoundé, Cameroon.

## Abstract

**Objective:** To evaluate the impact of the COVID-19 pandemic on the delivery of cardiology services in Africa.

**Design:** Cross-sectional e-survey study.

**Setting:** African countries

**Participants:** Cardiologists

**Primary and Secondary outcomes measures:** The primary outcome was the change in service delivery in African cardiology units during the on-going COVID-19 pandemic. The secondary outcomes were the satisfaction of cardiologists with regards to the workload and factors associated with this satisfaction.

**Results:** There was a significant reduction in working time and the number of patients consulted by week during this pandemic (p<0.001). In general, there was a decrease in the overall activities in cardiovascular care delivery. The majority of cardiology services (76.5%) and consulting programs (85%) were adjusted to the pandemic. Only half of the participants were satisfied with their workload. Reconfiguration of the consultation schedule was associated with a reduced satisfaction of participants (p=0.02).

**Conclusions:** COVID-19 is associated with an overall reduction in cardiology services rendered in Africa. Since the cardiovascular burdens continue to increase in this part of the World and the risk of cardiovascular complications linked to SARS COV2 remains unchanged cardiology, departments in Africa should anticipate a significant surge of cardiology services demanded by patients after the COVID-19 pandemic.

**Strengths and limitations of this study:**

- The study is one of the first African studies to report the impact of the COVID-19 pandemic on the delivery of cardiology services which are very important for Africans given the high prevalence of cardiovascular diseases in this continent.
- The multinational design of the study leading to the inclusion of 14 African countries makes the results generalizable to the entire African.
- The cross-sectional design of the study represents a major limitation as it remains impossible to either infer causality or untangle bi-directional relationships between the reduction of the delivery in cardiology services and the pandemic or participants’ satisfaction.
- Also, the e-survey was drawn in English and this might have restricted the participation by some non-English African respondents due to the language barrier. Hence, perhaps contributing to the relatively small sample size of the study.

## Introduction

Coronavirus Disease-2019 (COVID-19) is an unprecedented global health crisis. All sectors in all nations are affected by this pandemic. It is worth mentioning that health care systems bear the greatest burden in all societies (1). Since the start of the COVID-19 pandemic, health care delivery was reconfigured to meet up with health care demands while concomitantly making sure patients and health personnel do not get infected by this highly contagious and potentially lethal disease. Surgical departments were the most affected and consequently, they were the first to adjust the delivery of their services when the pandemic started (2). Next, Cardiology departments followed due to COVID-19 complications which are mainly cardiovascular such as thromboembolic disease, myocarditis, cardiac arrhythmia and ischemic heart diseases. Indeed, there is a strong relationship between COVID-19 crisis and cardiovascular burden which have prompted health policies to pay particular attention to the delivery of cardiology services (3-5). The European Society of Cardiology has published recent guidelines for the management of patients during this worldwide crisis (4). These guidelines include : a strategy for diagnosing of SARS-COV2, the viral pathogen of COVID-19, in cardiology units, triage of patients, teleconsultation, categorization of the emergency/urgency of invasive cardiovascular procedures, amendment of treatment protocols, recommendation to dispose of Personal Protective Equipment (PPE) to health care providers(4). At the time of this write-up, little was known about the impact of COVID-19 on the delivery of cardiology services. Fersla and colleagues in the UK found a 50% drop in cardiology departments’ admissions and a significant reduction in overall cardiology activities including outpatient consultations, diagnostic testing’s, procedures and cardiology community services such as cardiac rehabilitation (6). With an extensive literature search, to the best of our knowledge, the impact of COVID-19 on the delivery of cardiology services has not been reported on the African continent. Recent high-quality evidence showed Africa has the highest burden of cardiovascular diseases (CVD) due to the global epidemiological transition from infectious to non-communicable diseases before the outbreak of COVID-19 (7). Consequently, studying the impact of this pandemic on the delivery of cardiology services in Africa is crucial as COVID-19-related cardiovascular complications may worsen the CVD burden in this already highly affected and resource-limited continent. We aimed to evaluate the impact of COVID-19 on the delivery of cardiology services in Africa.

## Methods

### Study design/ Setting/ Participants

This was a cross-sectional study conducted using an online survey designed on Google Forms. The link was disseminated to several online networks of African cardiologists, scientific cardiology societies like the Cameroon Society of Cardiology, African Heart Rhythm Association, Pan-African Society of Cardiology, as well as the WhatsApp groups of Associations all African cardiology residents and interns. The questions were both mixed with closed and Likert scale items. The survey was piloted among cardiologists and senior’s cardiology residents/interns working and training respectively in Africa. The Survey had a total of 25 questions concerning demographic data : age, gender and nationality; changes in working time/number of patients consulted; change in the number of cardiovascular investigations requested by participants : 12 Lead Electrocardiography (ECG), ECG Holter, Ambulatory Blood Pressure Monitoring (ABPM), trans-thoracic and trans-esophageal echocardiography, stress electrocardiography, coronarography, ablations, pacing, electrophysiology; changes in patterns of CVD admissions the during COVID-19 pandemic; the level of satisfaction of participants (good, poor and unchanged). The details of the Google Form questions can be accessed at https://docs.google.com/forms/d/1A3SBAqwyKvIOhv-rI7LArBVqvUNIDulBmBct9pkQWv0/edit. To improve the rate of participation, reminder messages were sent to all the networks every 3 days. Data were collected over two week period spanning from the 05 to 20^th^ July 2020.

### Endpoints

#### Primary outcome

The change in activities in cardiology units during the COVID-19 pandemic.

#### Secondary outcomes

The satisfaction of participants with regards to workload activity and the factors associated with satisfaction

### Statistical methods

Analyses were done using Epi-Info version 7 Software. All data were expressed as proportions and frequencies. Bivariates analyses mainly Chi-square and Fisher Exact tests were used to find the factors associated with the satisfaction of the workload activity. A *p*-value<0.05 was considered as statistically significant.

## Results

### Characteristics of study participants

A total of 60 participants were recruited. The male gender was predominant (71.67%). The population was relatively young with 55% of them aged between 26-35 years. More than half (51.67%) were cardiologists and the rest were the seniors residents/interns in cardiology. A total of 14 African Countries were represented and the majority of participants worked in Cameroon (40%), followed by Ivory Coast (13%) and Senegal (10%). **See Figure 1**.

**Figure 1:**
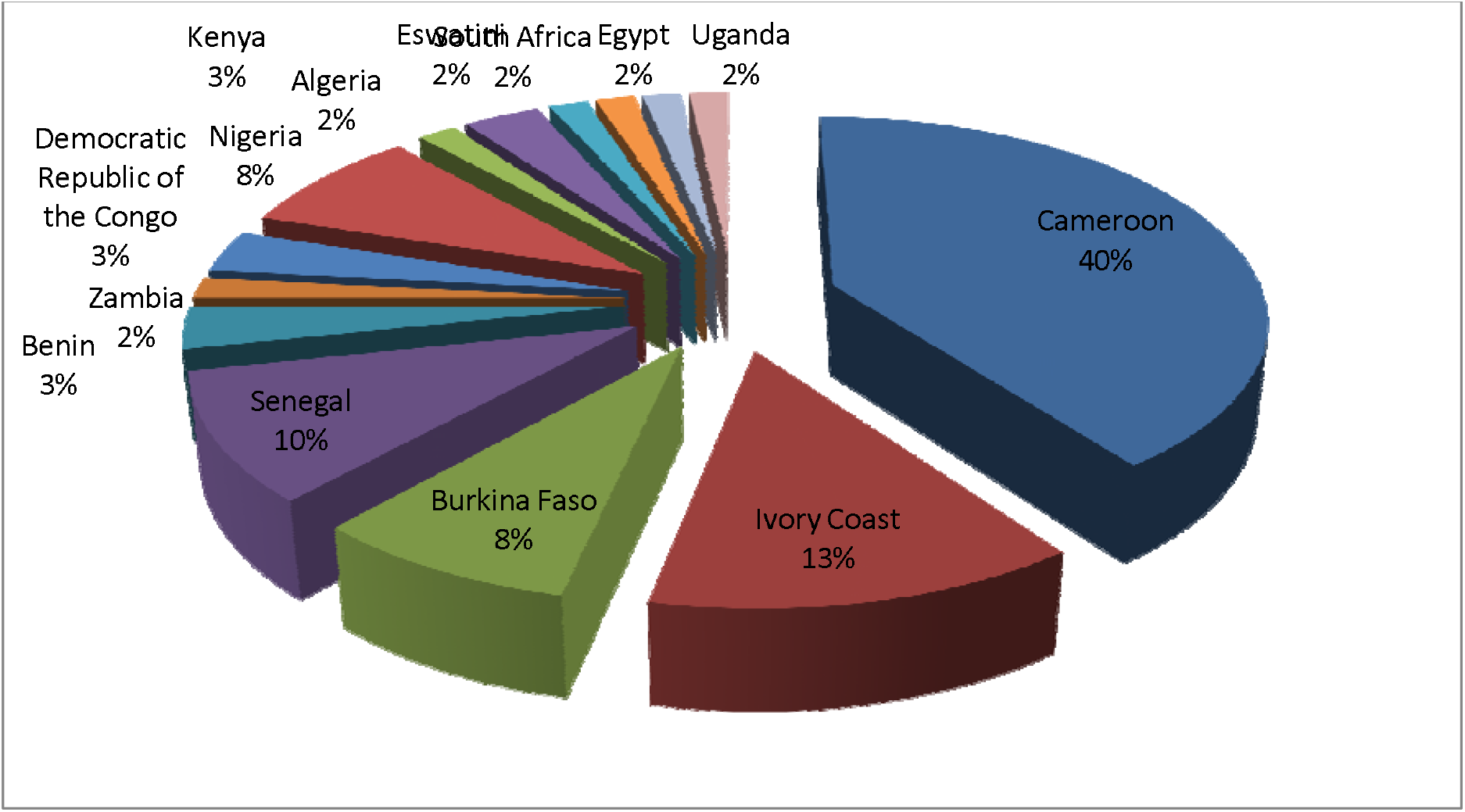
Distribution of the population according to the country.

### Outcome data

#### Changes in the delivery of cardiology services during COVID-19 pandemic

##### Changes in working time/number of patients consulted

There was a significant reduction in working time during the pandemic study period (p<0.001). Before the COVID-19 pandemic, the majority of participants (62%) had at least 30 hours per week of outpatient cardiology consultations. During the pandemic, only one-fifth (22%) of participants had 30 hours of consultations (**See table 1**).

**Table 1:** Working time during the COVID-19 pandemic.

| Working time<br>(hours per week) | Before COVID-19<br>N (%) | During COVID-19<br>N(%) | P value |
| --- | --- | --- | --- |
| <10 | 6 (10) | 13 (21.7) | <0.001 |
| 10 - 19 | 3 (5) | 15 (25) |  |
| 20 - 29 | 14 (23.3) | 19 (31.7) |  |
| ≥30 | 37 (61.7) | 13 (21.7) |  |

There was a significant reduction in the number of patients consulted per week during this pandemic (p<0.001). The majority of participants (33%) consulted about 20 to 30 patients per week before COVID-19, whereas during the pandemic most of them (38%) consulted less than 10 patients per week (**See table 2**).

**Table 2:** Number of patients consulting per week during COVID-19.

| Number of patients<br>consulted<br>(per week) | Before COVID-19<br>N (%) | During COVID-19<br>N (%) | P value |
| --- | --- | --- | --- |
| <10 | 2 (3.3) | 23 (38.3) | <0.001 |
| 10 - 19 | 14 (23.3) | 20 (33.3) |  |
| 20 – 29 | 20 (33.3) | 10 (16.7) |  |
| 30 – 39 | 10 (16.7) | 03 (5) |  |
| 40 – 49 | 03 (5) | 01 (1.7) |  |
| ≥50 | 11 (18.3) | 03 (5) |  |

##### Changes in the activities and clinical issue during COVID-19 pandemic

In general, there was a decrease in the global activities in cardiology services rendered to patients. The majority of cardiology services (76.5%) and consulting programs (85%) were reconfigured to meet up with the exigencies of the pandemic. The use of teleconsultation was also increased in daily practice (53.4%). The death rate was unchanged in the majority of the cases (55%). Diagnostic tests for COVID-19 were low (26.7%) and only one-third of participants received Personal Protective Equipment (PPE) regularly. Almost half (40%) of patients reported an increased level of cardiovascular complications during this pandemic. The most frequent cardiovascular disease during the pandemic study period was thromboembolic disease (41.7%) followed by cardiac arrhythmia (10%). Only half of the participants were satisfied by their workload during the pandemic. The change in activities and clinical issues in cardiology units during this pandemic are summarized in **table 3**.

**Table 3:** Changes in activities and clinical issue in cardiology units during the pandemic.

| **Activities/clinical issue** | **Frequency**<br>**N (%)** |
| --- | --- |
| **Turnover of patients** |  |
| *Decreased* | 49 (81.7) |
| *Increased* | 04 (6.7) |
| *Unchanged* | 07 (11.7) |
| **Modifications of the therapeutic protocol** | 28 (46.7) |
| **Changes in the death rate** |  |
| *Decreased* | 08 (13.3) |
| *Increased* | 19 (31.7) |
| *Unchanged* | 33 (55) |
| **Routine use of teleconsultation** |  |
| *Decreased* | 02 (3.4) |
| *Increased* | 31 (53.4) |
| *Unchanged* | 24 (41.4) |
| *Not used* | 01 (1.7) |
| **Rate of performance of basic cardiovascular exploration** |  |
| *Decreased* | 49 (81.7) |
| *Increased* | 4 (6.7) |
| *Unchanged* | 7 (11.7) |
| **Rate of performance of invasive explorations and therapies** |  |
| *Decreased* | 32 (53.3) |
| *Increased* | 02 (3.4) |
| *Unchanged* | 17 (28.3) |
| *Not performed* | 09 (15) |
| **Frequency of distribution of personal protective equipment** |  |
| *Never* | 04 (6.7) |
| *Rarely* | 14 (23.3) |
| *Regularly* | 23 (38.3) |
| *Always* | 11 (18.3) |
| **Rate of cardiovascular complications** |  |
| *Decreased* | 12 (20) |
| *Increased* | 24 (40) |
| *Unchanged* | 24 (40) |
| **Influenced by some cardiovascular diseases/ complications in daily practices** |  |
| Thromboembolic disease | 25 (41.7) |
| Arrhythmia | 06 (10) |
| Ischemic heart disease | 01 (1.7) |
| Heart failure | 04 (6.7) |
| Myocarditis | 03 (5) |
| Others | 04 (6.7) |
| **Satisfaction of workload in cardiology units during pandemic** | 32 (53.3) |

##### Factors associated with satisfaction of workload in cardiology units during COVID-19 pandemic

The satisfaction of the workload in cardiology units by participants was influenced by adjustment of the outpatient consultation schedule. There was an 89 % reduction (p=0.02) of satisfaction when the consultation program was adjusted during this pandemic. **See table 4**.

**Table 4:** Determinants of workload satisfaction in the cardiology units.

| Variables | Satisfaction of workload |  |  |  |
| --- | --- | --- | --- | --- |
|  | Yes<br>N= 32 | No<br>N= 28 | OR (IC 95) | P value |
| **Age**<br><35<br>≥35 | 19<br>13 | 15<br>13 | 1.27 (0.45-3.5) | 0.65 |
| **Gender**<br>*Female*<br>*Male* | 9<br>23 | 8<br>20 | 0.97 (0.31-3.01) | 0.97 |
| **Qualification**<br>*Cardiologist*<br>*Resident/Intern* | 15<br>22 | 15<br>13 | 0.59 (0.21-1.59) | 0.29 |
| **Reconfiguration of the consultation**<br>Yes<br>No | 24<br>8 | 27<br>1 | **0.11 (0.01-0.98)** | **0.02** |
| **Re-arrangement of the service**<br>Yes<br>No | 17<br>15 | 14<br>14 | 1.13 (0.14-3.1) | 0.8 |
| **Routine test for COVID-19**<br>Yes<br>No | 10<br>22 | 6<br>12 | 0.90 (0.26-3.1) | 0.87 |

## Discussion

COVID-19 is responsible for a tremendous crisis in several healthcare systems across the world (1). In this report, we found that there was a significant reduction in the number of hours spent at work and the number of patients consulted per week by cardiologists and residents/interns in cardiology during this pandemic. In general, there was a drastic drop in the overall activities carried out in cardiology units. The majority of cardiology services (76.5%) and outpatient cardiology consultations (85%) were reconfigured to health care demands of the pandemic. Only half of the participants were satisfied with their workload during the pandemic. The reconfiguration of the consultation program was statistically significantly associated with the reduction of participants’ satisfaction during this pandemic.

The reduction of admissions in cardiology departments is probably secondary to the fear of contracting COVID-19, known to be highly contagious (8). Another explanation for the reduction in consultations and admission in cardiology services are socio-cultural beliefs. Patients are scared that they will be quarantined and in the event of death, they will be buried immediately without them receiving proper last respect from their loved ones. In effect, the way the deceased patients are mourned in Africa is different from that in Western countries (9). Reconfiguration of the services mainly by putting in place a triage zone, medical or surgical staff by teleconsultation, distribution of PPE have been adopted by many healthcare systems to reduce the rate of infection among healthcare providers in general, which is reported to be high (10-12). It is worth mentioning that because this pandemic is unprecedented and its impact on the delivery of cardiology services have not been much explored, data on the relationship between COVID-19 and cardiology services are scarce. Similar to Fersla et al in the United Kingdom, we found a significant drop in cardiology units’ admissions and a significant reduction in the overall cardiology activities including outpatient clinics, investigations, procedures and community services such as cardiac rehabilitation (6). The new adaptation of the consultation programs was not appreciated by half of the clinicians, probably because it decreased their workload and consequently resulted in a reduction of their remuneration.

The major concern of this crisis is that, as the number of outpatient consultation reduced, the cardiovascular burdens continue to increase and the risk of SARS-COV2 to cause cardiovascular complications in COVID-19 patients remained unchanged (3). This could lead to a rebound in cardiovascular admissions and a marked increase in the workload of healthcare delivered after this pandemic. It is important to keep this in mind for better reconfiguration of services after COVID-19.

Lastly, although it was a multinational African survey, only 14 (26%) out of 54 African countries participated. This implies the cautious generalization of the study findings. However, the results of this pilot study were similar to trends seen abroad (6). Hence, the findings imply a scale-up in the delivery of cardiology services in Africa during and after the pandemic geared at better patient management COVID-19 patients. This cannot be overemphasized in a continent already bearing the highest-burden of CVDs and CVDs-related premature deaths (7).

## Conclusions

As observed in western countries, COVID-19 in Africa is associated with a reduction in the delivery of cardiology services. Since many patients no longer consult cardiology departments for CVD because of the fear of being infected by COVID-19 in African hospitals, these health facilities should be prepared to handle a massive influx of patients with CVD after the COVID-19 pandemic.

## Data Availability

Data could be access to https://docs.google.com/forms/d/1A3SBAqwyKvIOhv-rI7LArBVqvUNIDulBmBct9pkQWv0/edit.

## Author’s contribution

***Conception of the study:*** CNNG, MNT

***Data collection:*** CNNG, MNT, JNT, CN, VN,

***Manuscript writing:*** MNT, JNT, CN

***Revisions and approval of final manuscript:*** All the authors

***Supervision:*** AM, SK

## Acknowledgements

We acknowledge the Cameroon Society of Cardiology and the Africa Heart Rhythm Association, Pan-African Society of Cardiology for their great contribution to the survey.

